# Current applications of artificial intelligence for Fuchs endothelial corneal dystrophy: a systematic review

**DOI:** 10.1101/2024.11.16.24317435

**Authors:** Siyin Liu, Lynn Kandakji, Aleksander Stupnicki, Dayyanah Sumodhee, Marcello Leucci, Scott Hau, Shafi Balal, Arthur Okonkwo, Ismail Moghul, Bruce Allan, Dan Gore, Kirithika Muthusamy, Alison Hardcastle, Alice E Davidson, Petra Liskova, Nikolas Pontikos

## Abstract

**Purpose:** Fuchs Endothelial Corneal Dystrophy (FECD) is a common, age-related cause of visual impairment. This systematic review synthesizes evidence from the literature on Artificial Intelligence (AI) models developed for the diagnosis and management of FECD.

**Methods:** We conducted a systematic literature search in MEDLINE, PubMed, Web of Science, and Scopus from January 1, 2000, to June 31, 2024. Full-text studies utilizing AI for various clinical contexts of FECD management were included. Data extraction covered model development, predicted outcomes, validation, and model performance metrics. We graded included studies using the Quality Assessment of Diagnostic Accuracies Studies 2 tool. This review adheres to the PRISMA (Preferred Reporting Items for Systematic Reviews and Meta-Analyses) recommendations.

**Results:** Nineteen studies were analyzed. Primary AI algorithms applied in FECD diagnosis and management included neural network architectures specialized for computer vision, utilized on confocal or specular microscopy images, or anterior segment optical coherence tomography images. AI was employed in diverse clinical contexts, such as assessing corneal endothelium and edema, and predicting post-corneal transplantation graft detachment and survival. Despite many studies reporting promising model performance, a notable limitation was that only 3 studies performed external validation. Bias introduced by patient selection processes and experimental designs was evident in the included studies.

**Conclusions:** Despite the potential of AI algorithms to enhance FECD diagnosis and prognostication, further work is required to evaluate their real-world applicability and clinical utility.

**Translational Relevance:** This review offers critical insights for researchers, clinicians, and policymakers, aiding their understanding of existing AI research in FECD management and guiding future health service strategies.

## Introduction

Fuchs Endothelial Corneal Dystrophy (FECD) is a common, age-related cause of visual dysfunction estimated to affect 4-5% of individuals older than 50 years^1^. Characterized by the degeneration of the corneal endothelium, a critical monolayer of cells on the inner surface of the cornea, FECD leads to premature and progressive loss of these endothelial cells^2^. Corneal endothelial cell density declines with age in FECD, eventually falling below the density of cells required to prevent endothelial decompensation that leads to corneal edema, resulting in loss of corneal transparency, glare and impaired vision. Corneal transplantation becomes the sole definitive treatment option at this stage, with FECD being the primary indication for posterior lamellar keratoplasty in the United Kingdom^3^.

Historically, grading the severity of FECD involved quantifying guttata and detecting corneal edema through slit lamp biomicroscopic examination^4^. However, this approach has limited clinical utility due to significant interobserver variability in counting and describing guttata. Although end-stage corneal edema is evident on clinical examination, subclinical edema, by definition, is undetectable to the naked eye. Corneal densitometry and specific tomographic features derived from Scheimpflug camera tomography have been suggested as alternative methods for detecting and quantifying corneal edema^5,6^. Anterior segment OCT (AS-OCT) offers high-resolution cross-sectional scans of the cornea, but subtle localized swelling might be challenging to detect by humans from these images^7^. Current criteria for diagnosing and classifying the severity of FECD have been summarized recently^8^.

The rapid advances in artificial intelligence (AI) have sparked transformative changes across various medical fields, including ophthalmology. The ophthalmic community is uniquely positioned to harness AI strategies due to the widespread use of imaging tools in clinical practice. Over the past decade, AI has made significant strides in screening and detection of retinal diseases like diabetic retinopathy, age-related macular degeneration and inherited retinal diseases^9–12^. This progress has been made possible by the availability of large and diverse labelled datasets. More recently, promising results have also been reported in anterior segment conditions, namely keratoconus^13^ and refractive surgery screening^14^. Despite the prevalence of FECD, application of AI in this area has been relatively limited.

This systematic review aims to synthesize and evaluate the quality of reporting, risk of bias, the different types of AI algorithms and their performance for FECD. Our goal is to accurately portray the current state of development, identifying the opportunities and challenges to implementing AI in routine clinical practice.

## Methods

### Search Strategy

We conducted a literature review of AI applied to the diagnosis and management of FECD published between January 1, 2000, and August 31, 2023. The PRISMA (Preferred Reporting Items for Systematic Reviews and Meta-Analyses) Statement 2020 criteria^15^ were followed by searching bibliographic databases (PubMed, MEDLINE, EMBASE, Web of Science, Scopus, Cochrane Library, IEEE Explorer, Digital Library) using keyword search on their title, abstract, and keywords. It has been registered *a priori* in the International Prospective Register of Systematic Reviews (PROSPERO ID: CRD42023454518). The search term included a combination of the following terms: ("Fuchs endothelial corneal dystrophy” OR “Fuchs dystrophy” OR “endothelial dystrophy” OR “guttata”) AND (“algorithm” OR “artificial intelligence” OR “machine learning” OR “deep learning”) AND (*“detection” OR “diagnosis” OR “grading” OR “classification” OR “segmentation”).*In each database, the same search terms were used and a filter was set to include human studies and exclude animal studies. Relevant studies were also identified via a search in Google Scholar or from the bibliographies of the included studies.

### Eligibility Criteria

Studies were included if they investigated the application of AI algorithms for the detection of FECD, classification of FECD disease severity, identifying corneal edema, assessment of the corneal endothelium in FECD patients, diagnosis of graft detachment, and prediction of post-corneal transplantation outcomes. The studies should have reported the performance of the AI model employed using metrics appropriate for the task, such as sensitivity and specificity for classification or the Sørensen–Dice coefficient for segmentation. The performance metric should derive from a validation or test set separate from the training data set. Finally, the full-text article must be available, and only papers published in English were considered. Studies were excluded if they were not conducted on humans, focused on guttata and/or corneal edema caused by factors other than FECD, or involved eyes complicated by other ophthalmic conditions. Additionally, this review excluded articles classified as case reports, reviews, comments, qualitative studies or protocols.

### Data Synthesis

The PRISMA flowchart (**Figure. 1**) provides an overview of the study search and selection process. Three reviewers (SL, LK and AS) screened the initial results based on the inclusion and exclusion criteria. Disagreements in meeting the inclusion or exclusion criteria were resolved by discussion. Following this, data were extracted from studies that met the eligibility criteria: author and year, title, country, age, gender, number of eyes for each group, clinical utility, validation details, input data, algorithm, classification groups, performance metrics appropriate for the task (e.g. sensitivity, specificity, accuracy, precision, area under the receiver operating characteristic (ROC) curve [AUC], dice coefficient). Due to the heterogeneity in the described algorithms, study methodologies, and reported metrics, meta-analysis was considered inappropriate and was not performed.

**Figure 1.**
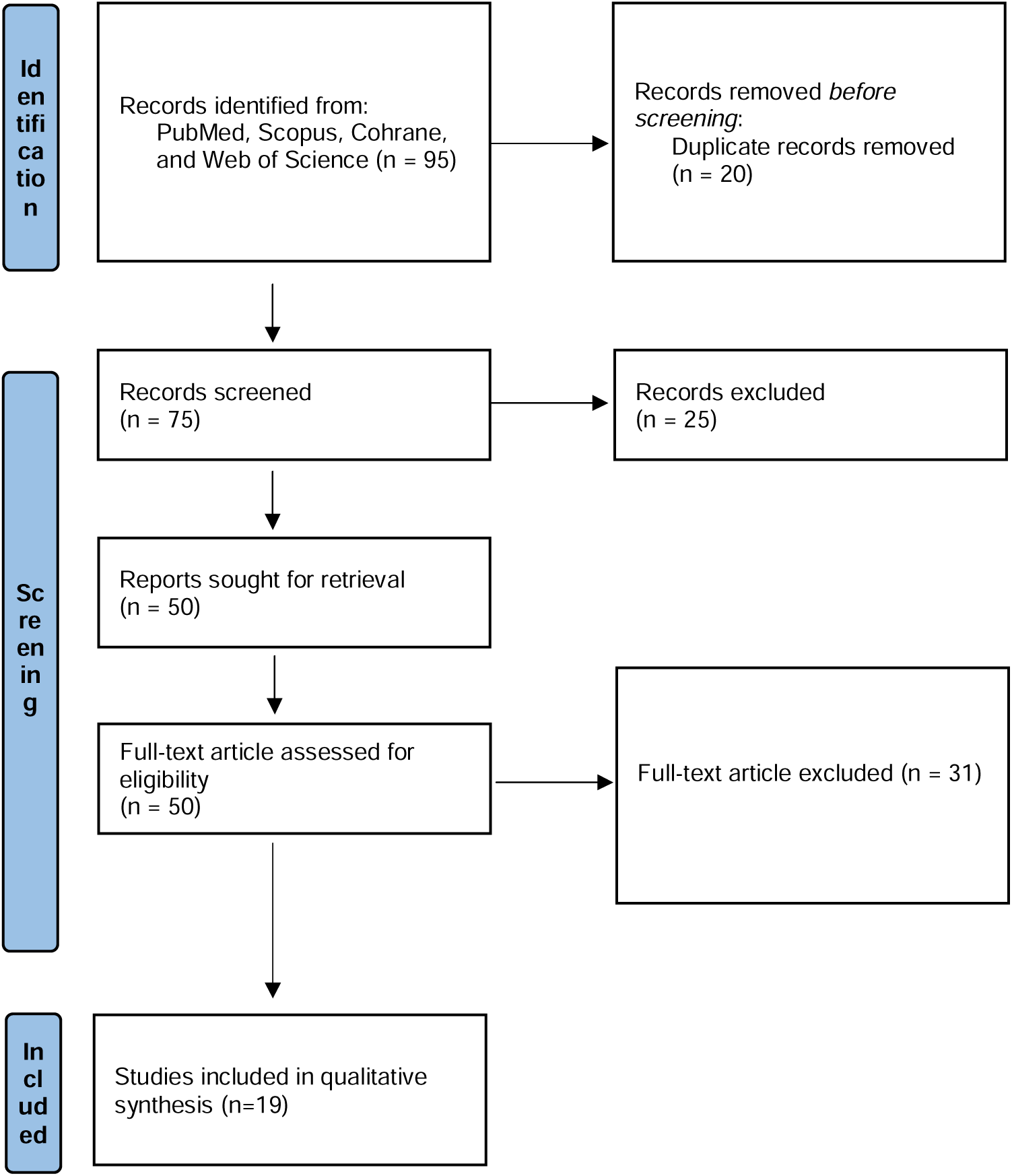
PRISMA (Preferred Reporting Items for Systematic Reviews and Meta-Analyses) flow diagram displaying filtering steps taken to accept or exclude studies in the systematic review.

### Bias Assessment

When evaluating bias across the included studies, we employed the Quality Assessment of Diagnostic Accuracy Studies-2 (QUADAS-2)^16^, a tool designed to critically appraise the methodological quality and potential bias in studies that evaluate the diagnostic accuracy of medical tests, which assesses four key domains: patient selection, index test, reference standard, and flow/timing. Three independent reviewers (SL, LK, and AS) conducted the assessments, with each study reviewed by at least two reviewers for a reliable assessment. The correlation between adherence to QUADAS-2, measured by the percentage of “Yes” responses, and both the year of publication and the latest available journal impact factor (as reported by the World of Science), was analysed using a linear regression model in R Statistical Software (R Foundation for Statistical Computing, Austria).

## Results

### Literature Search

The study selection process is outlined in **Figure 1**. The literature search initially identified 76 studies. Following the removal of 20 duplicate records, 25 studies were excluded based on relevance, including the exclusion of animal studies and those that did not investigate clinical applications to FECD, leaving 50 studies for full-text review. A detailed breakdown of the reasons for including or excluding each article is provided in **Supplementary Table 1.** Ultimately, 19 articles were included in our systematic review. **Table 1** outlines the datasets and the developed AI pipelines used in the individual studies.

**Table 1:**
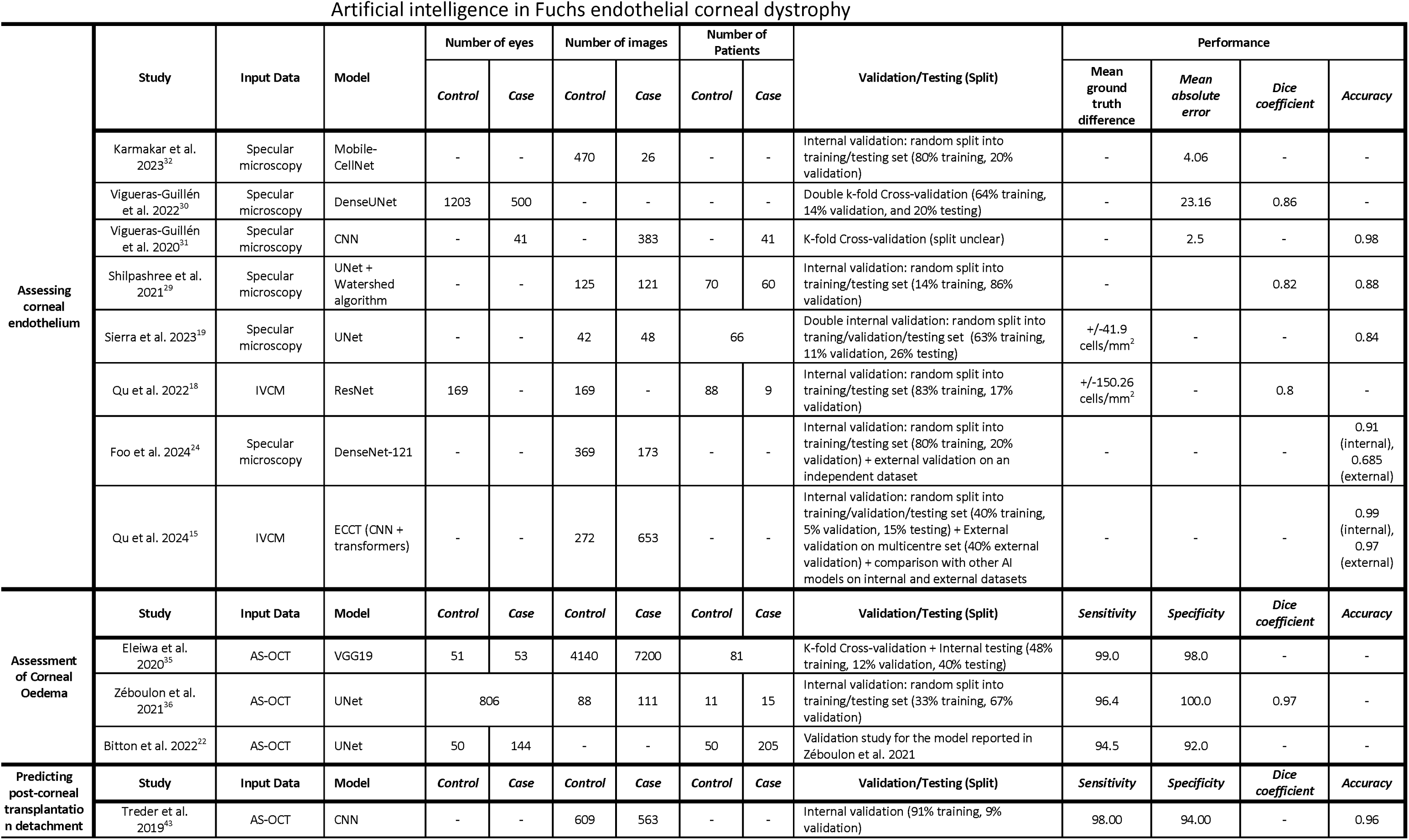
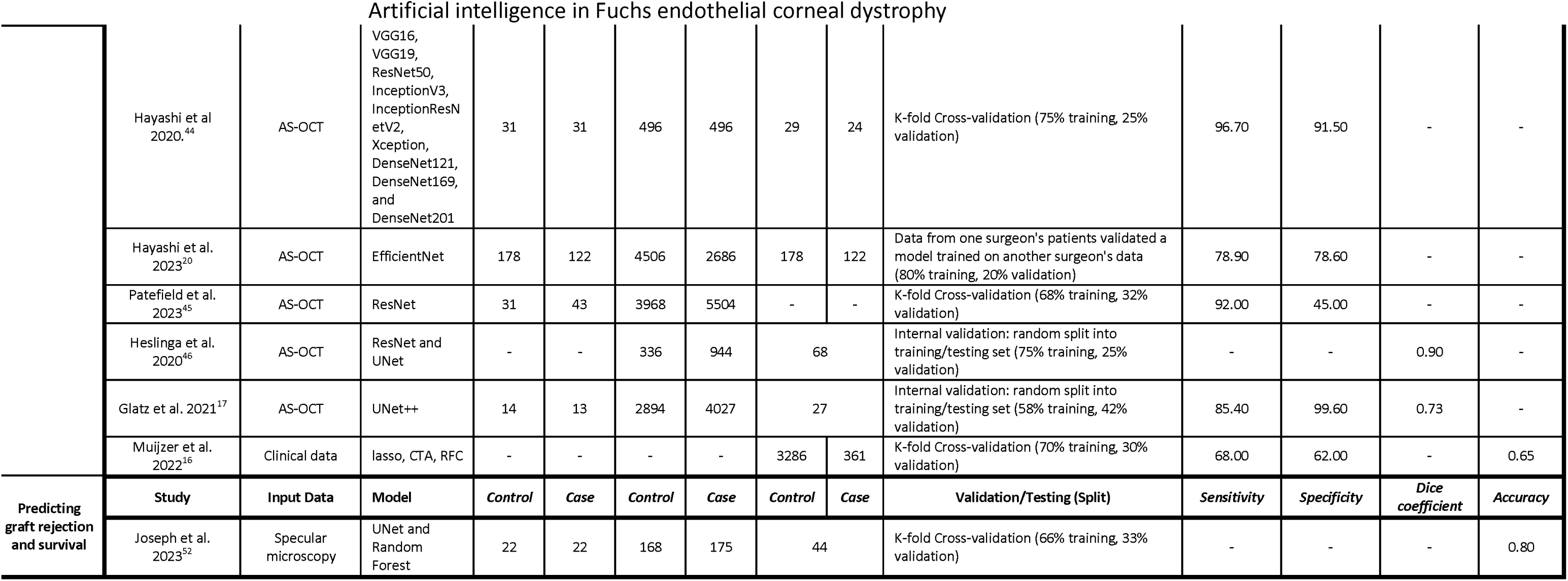
Summary of the published studies that included the use of artificial intelligence for the assessment, diagnosis, and prognostication of Fuchs Endothelial Corneal Dystrophy (FECD). Bland Altmann plot given for average difference from ground truth. Abbreviation: FECD, Fuchs endothelial corneal dystrophy;AS-OCT, anterior segment-optical coherent tomography; DMEK, Descemet membrane endothelial keratoplasty; PBK, Pseudophakic bullous keratopathy; PK, Penetrating keratoplasty.

### Input Datasets for Model Development

The datasets utilized in these studies had a median sample size of 925 images (range: 30-11340 images), with a median ratio of 1.09 (range: 0.05-3.14) FECD cases per control. Primarily, algorithms were trained using single-center data (n=15, 78.9%), with one using multicenter (n=1, 5.3%)^17^ and one study using national registry (n=1, 5.3%)^18^ datasets. In 3 studies, the source of patients could not be determined^19–21^. None of the studies utilized public datasets for model development.

### Imaging Modalities

Except for the one study utilizing national registry-derived clinical datasets for training algorithms predicting graft detachment^18^, most of the models included in the review were trained using raw corneal images. Predominantly, cross-sectional images of the cornea were acquired via AS-OCT, which served as the primary imaging modality for model development. These images were generated from various devices such as Envisu (Bioptigen, USA), Spectralis (Heidelberg Engineering, Germany), CASIA (Tomey Corp, Japan), and Avanti OCT (Optovue, USA). These systems can produce numerical indices measuring corneal metrics like thickness, keratometry, elevation, and aberrations, but the included studies train models using the raw images acquired instead of the processed indices. In addition, seven models focused on evaluating corneal endothelium, with seven employing images captured via specular microscopy (CellCheck XL [Konan Medical, Japan], SP-1P [Topcon Co, Japan], EM-3000 [Tomey Corp, Japan]), and one using confocal microscopy (HRT III Rostock Cornea Module [Heidelberg Engineering]).

### Algorithm Validation

Most of the studies included did not incorporate external validation (16/19); instead, they relied on internal validation methods such as k-fold cross-validation (n = 7) or random partitioning of the original dataset into training and validation sets (n = 9). Among these, one study by Hayashi et al. stands out for training the model using patient data from one surgeon to predict post-transplant graft detachment requiring rebubbling, then validating it with data from another surgeon^22^. The authors reported an AUC of 87.5% when applying the trained algorithm to the second surgeon’s dataset. However, since the model’s performance on the original dataset used for training and validating the algorithm was not provided, comparisons to assess overfitting are not feasible.

To evaluate and augment a prior model developed by the same group^23^ in a pre-operative setting, Bitton et al. trained the algorithm using AS-OCT images derived from 50 normal corneas and 240 edematous corneas of pre-Descemet membrane endothelial keratoplasty (DMEK) patients^24^. These patients had indications for DMEK due to either FECD (144 eyes) or pseudophakic bullous keratopathy (96 eyes). The reported optimal ’edema fraction (EF)’ threshold, representing the ratio between pixels labelled as corneal edema and those representing normal cornea, was 0.143. At this threshold, the model had a sensitivity of 94.5%, specificity of 92%, and an AUC of 96%, roughly in line with the performance of their internal validation using the original dataset. While the model demonstrated promising performance, in some control cases, a significant proportion of normal corneas were labelled as edematous. This occurrence may reflect global signal differences unrelated to the presence or absence of corneal edema. Caution is warranted as convolutional neural network (CNN) models can be highly sensitive to subtle signal differences that might not be discernible to the human eye^25^.

Recent studies have increasingly utilized external validation. For instance, Qu et al. used a multicenter dataset with images from seven hospitals, while Foo et al. employed an unspecified independent external validation set ^17,26^. Both studies observed a decline in performance, with Qu et al.’s algorithm showing a more pronounced drop. This performance reduction underscores a generalization gap and highlights the limitations of clinical applicability in real-world settings. Retrospective training datasets often undergo extensive filtering and cleaning, which may not accurately represent the variability of real-world data, leading to overfitting and diminished performance on external datasets.

### Clinical context, algorithms, and model performance

The primary AI algorithms applied in FECD diagnosis and management encompass neural network architectures specialized for computer vision, i.e., image classification or segmentation tasks. Muijzer et al. pursued regression and decision tree-based approaches for identifying risk factors for graft detachment^18^. This subsection is structured according to the clinical contexts where these AI algorithms were applied. Figure 2 is a summary diagram delineating the pertinent models utilized across different clinical scenarios.

**Figure 2.**
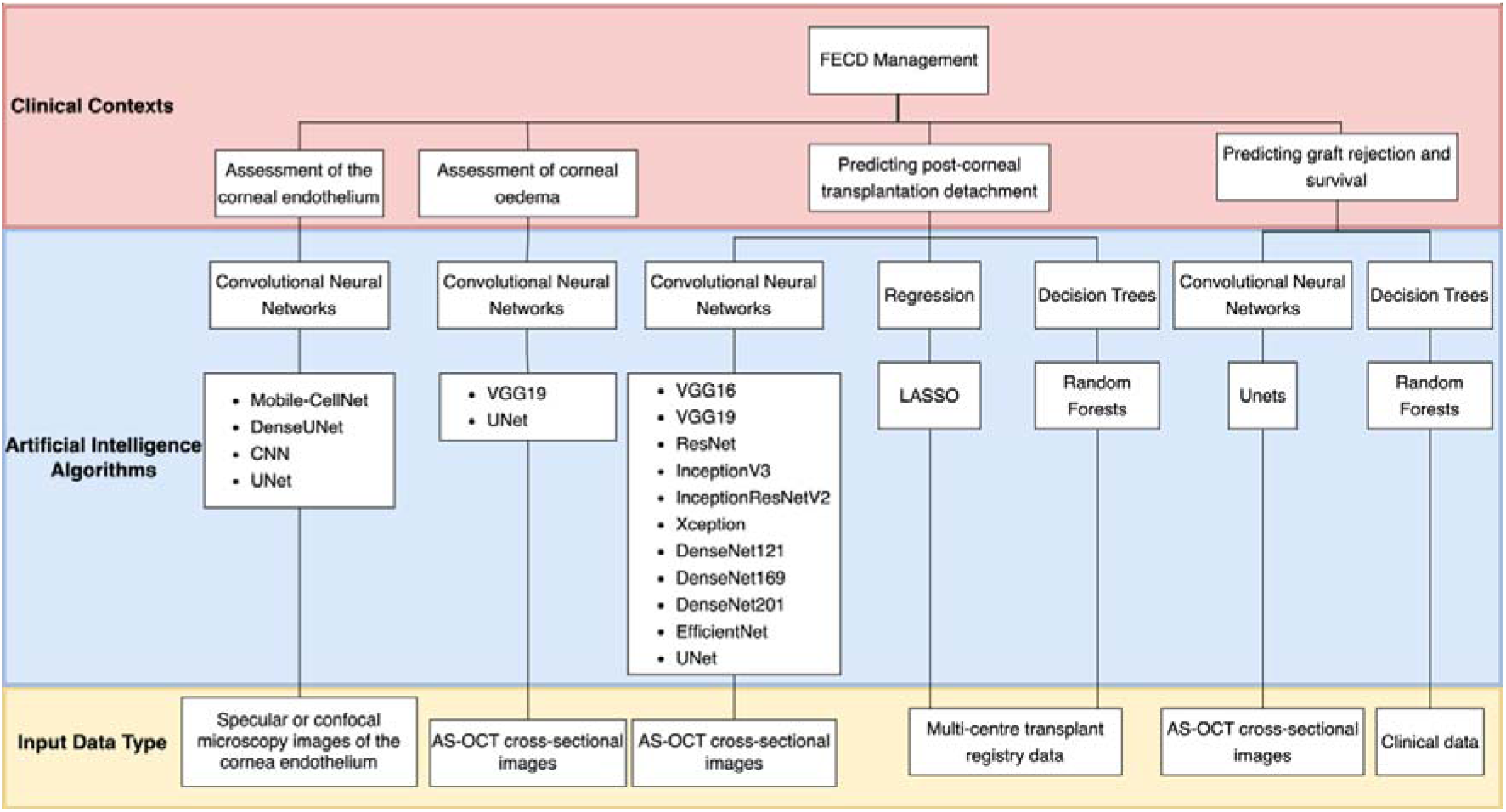
Summary diagram of the relevant AI approaches used across various clinical contexts of FECD diagnosis and management. Abbreviation: FECD, Fuchs endothelial corneal dystrophy; LASSO, least absolute shrinkage and selection operator; AS-OCT, anterior segment-optical coherent tomography.

#### Assessment of corneal endothelium

The endothelium is critical in evaluating FECD due to specific morphological patterns that can help diagnose and stage disease. Imaging of the endothelium can be achieved using various technologies such as specular microscopy and in vivo confocal microscopy (IVCM; Figure 3)^27^. These technologies allow for semi-manual or automated quantification of endothelial cell density (ECD) and morphology. Automated corneal endothelial cell segmentation has long relied on marker-driven watershed segmentation, but this method is prone to both under- and over-segmentation, especially in areas with low signal-noise ratio^28,29^. AI approaches have been applied to segment corneal endothelial cells more accurately ^30^. AI brings a significant advantage by adapting to variability in cell morphology and image quality, providing more precise segmentation where traditional methods fall short. Additionally, it can continuously improve through learning, allowing it to handle complex patterns and subtle changes that can be otherwise missed.

**Figure 3.**
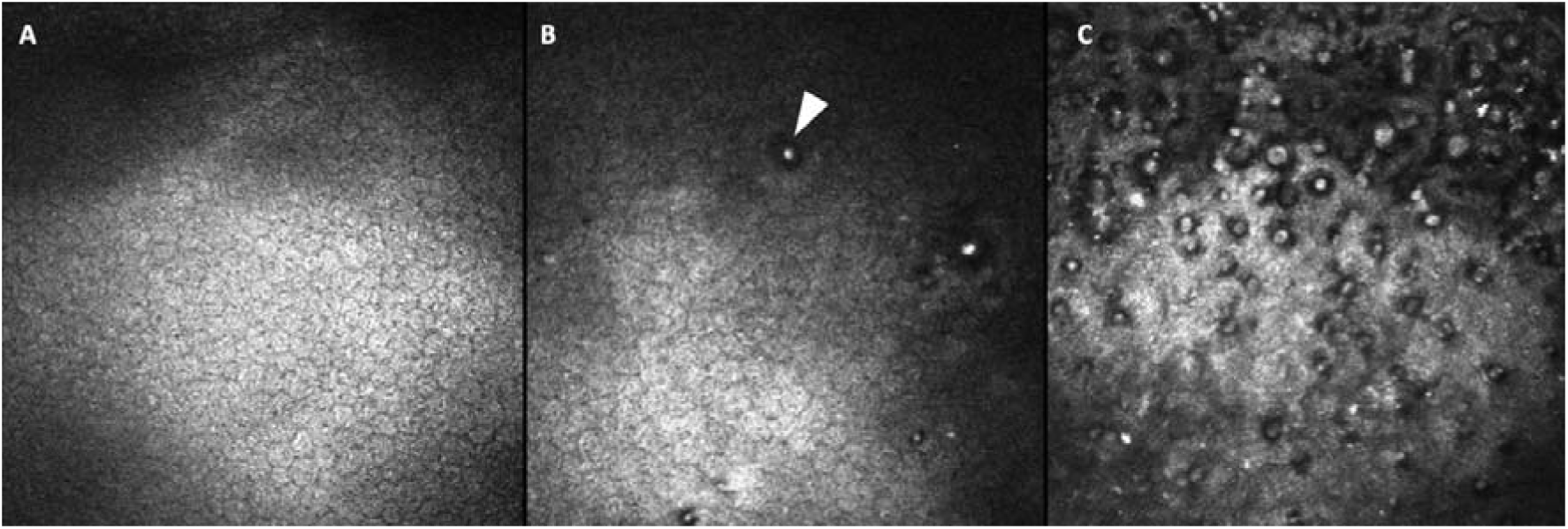
Slit-scanning in vivo confocal microscopy images of the corneal endothelium: A) Image of a healthy control cornea, showing normal endothelial cell morphology without signs of FECD. B) Image depicting isolated guttata, characterized by dark, round bodies with central white hyperreflectivity. C) Increased confluency of guttata, illustrating a more extensive presence of these structures.

Several studies have employed U-Net-based architectures for segmenting corneal endothelial cells from specular microscopy images. Shilpashree et al. used a modified U-Net with Watershed post-processing to resolve merged cell borders in images of FECD patients and healthy subjects. Their model yielded 87.90% accuracy in correctly segmenting the percentage of pixels in the image, an F1 score of 82.27%, and an AUC of 96.70% for recognizing the cell borders^31^. Sierra et al. approached the problem as a regression task of cell and gutta-signed distance maps instead of pixel-level classification, which converged faster and yielded 83.79% accuracy in guttae identification ^21^. These studies demonstrate the effectiveness of U-Net variants in analyzing corneal endothelial images in FECD.

Vigueras-Guillén et al. proposed a novel deep-learning method using Dense U-Net for segmenting specular microscopy images of the corneal endothelium with guttae^32^, incorporating an attention mechanism called *feedback non-local attention* (fNLA) to infer the cell edges in occluded areas. This binary segmentation enabled the estimation of ECD, coefficient of variation (CV), and hexagonality (HEX). Compared to manually segmented ground truth of 1203 images, they found that Dense U-Net with fNLA produced the lowest error, with a mean absolute error of 23.16 cells/mm² in ECD, 1.28% in CV, and 3.13% in HEX, which was 3-6 times smaller than the manufacturer’s (Topcon) built-in software. Vigueras-Guillén et al. also utilized Dense U-Net to estimate corneal endothelium parameters from specular microscopy images acquired with a Topcon SP-1P microscope of eyes 1, 3, 6, and 12 months post ultrathin Descemet stripping automated endothelial keratoplasty (UT-DSAEK) ^33^.

The proposed method outperformed Topcon’s software, with a mean absolute error of 30.00 cells/mm² (vs 118.70 cells/mm²) for estimating ECD, 1.5% (vs 4.6%) for CV, and 3.0% (vs 10.3%) for HEX.

Karmakar et al. proposed a novel deep learning-based cell segmentation algorithm called Mobile-CellNet to estimate ECD in specular microscopy images^34^. The proposed algorithm uses two similar CNN-based image segmentation models working in parallel along with image postprocessing using classical image processing techniques to delineate the boundaries of the endothelial cells. When compared to the widely used U-Net architecture, Mobile-CellNet resulted in a mean absolute error of 4.06% for ECD on the test set, offering similar performance but greater computationally efficient.

Foo et al. developed two classification algorithms using DenseNet-121 to detect FECD and analyse corneal endothelium in specular microscopy images, respectively. The first model achieved an AUC of 0.96, 91% sensitivity, and 91% specificity on an internal dataset, but these metrics dropped to 0.77, 69%, and 68% on an external dataset, performing worse than expert manual grading. Their second model identified widefield specular microscopy images with ECD > 1000 cells/mm² in FECD eyes, achieving an AUC of 0.88, 79% sensitivity, and 78% specificity.

The Heidelberg Rostock Cornea Module, commonly used for IVCM in clinical settings, lacks automated tools for assessment of corneal endothelium, prompting Qu et al. to develop a fully automated segmentation and morphometric parameter estimation system for assessing central corneal endothelial cells from IVCM images ^20^. The automated system calculated various morphometric parameters, including ECD, CV in cell area, and percentage of HEX. The automated ECD were highly correlated with ECD measurements from Topcon’s specular microscope (*r* = 0.932), which also correlated highly with ground truths ECD derived from manual calculation (*r* = 0.818). Given IVCM’s ability to provide cell-level visualization of corneal structure and superior optical penetration even in the presence of edema, an automated morphometric system for analyzing IVCM images could, as a proof of concept, offer a more effective diagnostic measure than specular microscopy.

Qu et al. also developed an open-access system for recognizing IVCM images of various corneal endothelial diseases, including FECD, using the novel Enhanced Compact Convolutional Transformer (ECCT), which combines CNNs and transformers.^17^. When validated on a multicenter testing dataset from seven Chinese hospitals, the model achieved a sensitivity of 98.3%, specificity of 94.8%, and accuracy of 97% in recognizing FECD images, demonstrating comparable or superior performance to other robust AI models, including ResNet-34, EfficientNet-B5, DeiT-S, and Swin-T.

#### Assessment of corneal edema

The presence of corneal edema **(Figure 4)** signifies a crucial stage in the progression of FECD, indicating a decline in ECD below the threshold required for maintaining corneal dehydration (referred to as stromal deturgescence). Although corneal thickness is widely used in clinical settings to measure corneal edema, it can be misleading in naturally thin or thick corneas^35,36^. Thus far, three studies have developed algorithms for assessing corneal edema, of which one aimed to classify the disease into early and late stages^37^, whilst the other two quantified the severity of edema^24,38^.

**Figure 4.**
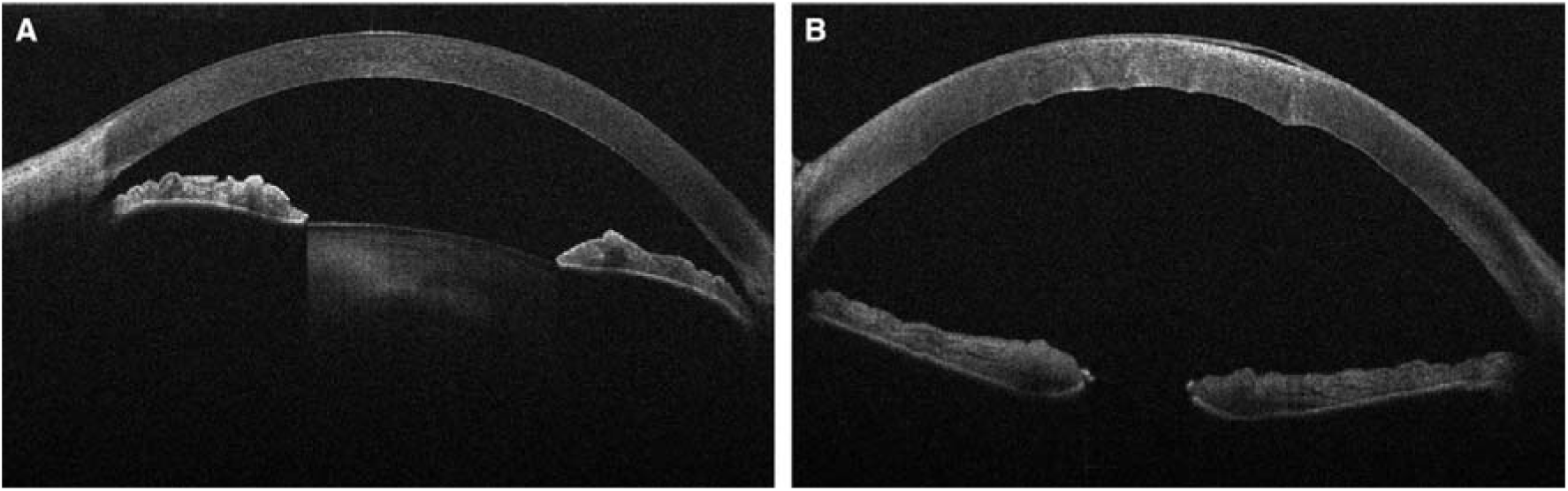
Cross-sectional AS-OCT images of the cornea in early-stage FECD without clinical signs of corneal oedema (**A**), and late-stage FECD showing increased central corneal thickness, epithelial bullae, and increased pixel intensity (backscatter), indicating corneal decompensation (**B**). *Abbreviation*: *FECD*, Fuchs endothelial corneal dystrophy; *AS-OCT*, anterior segment-optical coherent tomography.

CNNs have demonstrated the ability to make inferences from imaging data structures through deep learning^39^. Eleiwa et al. developed a deep learning model based on the Visual Geometry Group with 19 layers (VGG19) to classify AS-OCT images into normal, early-stage (defined as without clinical edema), and late-stage FECD (presence of clinically evident edema), with sensitivity of 91%, 100%, and 99%, specificity of 97%, 92%, and 98%, and AUCs of 0.997, 0.974, and 0.998, respectively^37^. However, this approach did not provide information on the precise location of edema on the images, which may limit its clinical application in detecting subclinical or focal edema.

Zéboulon et al. developed a U-Net-based^40^ segmentation model to identify corneal edema and derive the EF from AS-OCT images^38^. Trained on 111 scans of post-DMEK corneas with total corneal edema and 88 normal corneal scans, the model had Dice coefficients of 0.990 and 0.967 for normal and edema pixel predictions, respectively. With an optimal EF threshold of 0.068, the model had a sensitivity of 96.4%, specificity of 100%, and an AUC of 0.994 for distinguishing edematous from normal corneas^38^. Although this pilot study confirmed the potential of using a CNN model to detect corneal edema, the concurrent evaluation of the epithelium and stroma hindered its ability to detect mild stromal edema, which can be masked by a normal epithelium and lead to false negatives. The same research group later enhanced the model by incorporating additional layers to delineate the epithelium and stroma in the image analysis pipeline, enabling the new model to detect significant EF differences between normal corneas and those with mild edema^23^, a capability not achieved by the original model^38^.

#### Predicting post-corneal transplantation graft detachment

Graft detachment, a complications specific to DMEK^41^ (**Figure 5)**, has reported rates range from 4% to 34.6%^42^, with 0.2-75% of cases required post-operative re-bubbling^43^. AS-OCT is a superior modality for detecting subtle and shallow graft detachment compared to slit-lamp examination alone^44^. Seven studies investigated the potential of utilizing AI algorithms to recognize ^22,45–47^, quantify ^19,48^ and predict ^18^ post-corneal transplantation graft detachment.

**Figure 5.**
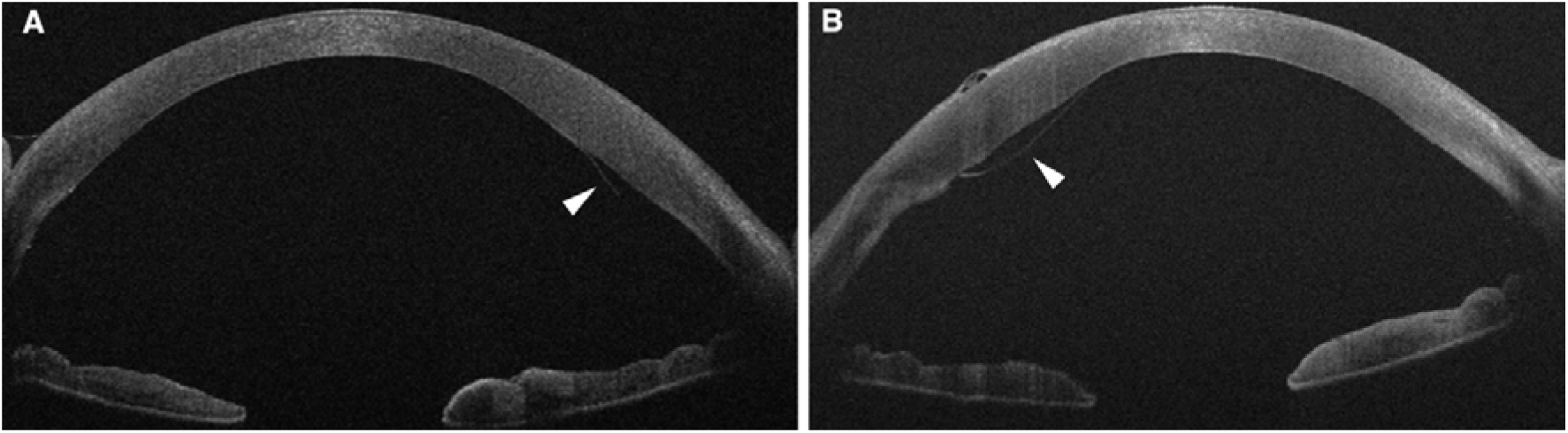
Cross-sectional AS-OCT images of the cornea showing a small partial graft detachment after DMEK (**A**), and a more extensive graft detachment (**B**). *Abbreviation*: *AS-OCT*, anterior segment-optical coherent tomography; *DMEK*, Descemet membrane endothelial keratoplasty.

Treder et al. utilized post-DMEK AS-OCT images to develop a CNN classifier based on a pre-trained algorithm for distinguishing corneas with attached grafts from those with graft detachment^45^. The input dataset comprised 1172 AS-OCT images (609: attached graft; 563: detached graft) from 111 eyes of 91 patients within the first 90 days postoperative period. The classifier achieved 98% sensitivity, 94% specificity, and 96% accuracy^45^. Irregular graft curvature, image artefacts, and flat or peripheral graft detachment were identified as potential sources of misclassification^45^.

Rather than simply classifying graft detachment, Hayashi et al. compared the performance of multiple deep-learning classification models (VGG16, VGG19, ResNet50, InceptionV3, InceptionResNetV2, Xception, DenseNet121, DenseNet169, and DenseNet201) to discriminate patients requiring re-bubbling after DMEK from those who did not^46^. Training these models with a dataset of 496 images from 31 eyes requiring re-bubbling and 496 images from 31 eyes not requiring re-bubbling, all operated by one surgeon, the VGG19 model exhibited the highest AUC of 0.964. The sensitivity and specificity of this model were reported as 0.967 and 0.915, respectively^46^. Subsequently, the same research group developed EfficientNet-based algorithm, trained on OCT images from patients operated on by one surgeon, which performed well in predicting the need for re-bubbling in surgeries by another surgeon, with AUC of 0.875, the Youden index of 0.214, and sensitivity and specificity of 78.9% and 78.6% respectively^22^.

The previously described graft detachment classification models are primarily based on AS-OCT scans obtained after DMEK surgery. However, an algorithm capable of predicting the risk of graft detachments before surgery could significantly optimize surgical care by serving as a pre-operative screening tool. Patefield et al. developed a novel Multiple-instance learning artificial intelligence (MIL-AI) model^49^ using ResNet and pre-operative AS-OCT images to distinguish between eyes with and without graft detachment after DMEK, with a sensitivity of 92%, a specificity of 45%, and an AUC of 0.63, outperforming the human labelling by ophthalmologists in sensitivity but demonstrating lower specificity^47^.

In contrast to binary classification approaches focusing on the presence or absence of graft detachment, deep learning-based segmentation techniques can quantify the length and location of the detachment. Heslinga et al.^48^ developed an automated method using AS-OCT images to locate and quantify graft detachment after DMEK. A ResNet-based^50^ model localized the scleral spur for anterior chamber measurements, followed by a U-Net architecture^40^ for semantic segmentation to identify image pixels corresponding to graft detachment. Trained and validated on 1280 AS-OCT images, of which 336 did not have graft detachment from 68 eyes post-DMEK, the model accurately estimated graft detachment lengths in 69% of cases, with a Dice score of 0.896^48^. However, the model’s performance in identifying large central detachments was limited, likely due to insufficient training samples and confusion with intraocular gas.

More recently, Glatz et al.^19^ developed a U-Net-based model to generate percentage detachment map and 3-dimensional volume representation of graft detachment after DMEK. Trained on 6912 manually labelled AS-OCT images from 27 eyes with FECD (including 14 eyes without graft detachment) after a median of 16 days post-DMEK, the model reported a Youden index of 0.850, a Dice coefficient of 0.729, a sensitivity of 0.854, and a specificity of 0.996^19^. In an application set of 107 eyes, the model’s outperformed slit-lamp examination by detecting detachments missed or underestimated by human specialists.

Rather than using an image-based computer vision system, Muijzer et al leveraged data from the Netherlands Organ Transplant Registry and employed three machine learning models, namely L1 regularized logistic regression with least absolute shrinkage and selection operator (lasso), classification tree algorithm (CTA), and random forest classification (RFC), to identify risk factors for graft detachment after posterior lamellar keratoplasty^18^. Key risk factors identified included undergoing a DMEK procedure (rather than DSAEK), prior graft failure, and using sulfur hexafluoride gas during surgery. The RFC model performed best with an AUC of 0.72, sensitivity of 0.68, and specificity of 0.62, compared to LASSO (AUC 0.70, sensitivity 0.70, specificity 0.65) and CTA (AUC 0.65, sensitivity 0.65, specificity 0.62)^18^.

#### Predicting graft rejection and survival

The assessment of ECD has been proposed as a predictor of graft rejection and survival^51–53^. Joseph et al. ^54^ developed a deep learning pipeline for identifying eyes at risk of graft rejection within 1 to 24 months after DSAEK. The authors used a pre-trained U-net^55^ model to segment endothelial cells and extract novel quantitative features relevant to cellular spatial arrangements and cell intensity values. These features were used to train RF and logistic regression models, with over 0.80 accuracy in predicting post-DMEK graft rejection. Key predictors for graft rejection included cell-graph spatial arrangement, intensity, and shape features. However, research in this area is limited partly because many endothelial rejection episodes are asymptomatic.

### Study quality

The risk of bias was assessed using the QUADAS-2 tool^16^. A wide range of adherence rates (percentage of questions answered “Yes” out of all the QUADAS-2 criteria) was observed (Median=54.6%, Interquartile range 45.5%-72.7%), indicating the variable quality of the included studies. In general, patient selection was found to have a high risk of bias in over half of the included studies (11/19, 55%) because the majority of these studies were case-control and did not use consecutive or random samples. A high risk of bias was introduced by the index test in 26% (5/19) of cases, mainly due to the result being interpreted with the knowledge of the results of the reference standards. However, a significant proportion of the articles did not specify this and were marked as ‘Unclear’. Reference standards for many classification and segmentation models consisted of manual interpretation and annotation by human experts. While no gold standards exist for these tasks, manual curation for reference results by trained specialists was considered a low risk of introducing bias. The individual assessment for each study is present in the **Supplementary Table 2**.

## Discussion

Our systematic review identified 19 peer-reviewed manuscripts that presented AI algorithms for diagnosing FECD, assessing corneal edema, evaluating corneal endothelium, and predicting post-corneal transplant outcomes. Notably, this review is the first systematic examination of AI applications in assessing FECD. Researchers have targeted various aspects of the patient journey, including diagnosis, severity grading, and prognostication, utilizing diverse imaging modalities and clinical data. Predominant areas of investigation involve predicting graft detachments post-posterior corneal transplantation and assessing corneal endothelium, followed by analyzing corneal edema and predicting graft survival.

In the current literature, the predominant input data are raw pixel-level corneal imaging obtained from either AS-OCT or specular microscopy. Conversely, developing models for the detection of keratoconus, another prevalent cornea disease, often relies on computed parameters such as keratometry and elevation data rather than direct corneal images ^13,56,57^. This difference in approach arises because FECD assessment heavily hinges on clinical examination, lacking validated numerical indices to differentiate disease stages, unlike keratoconus. Moreover, the input imaging data were produced by devices of different manufacturers. Despite sharing the same underlying technology and appearing similarly to human eyes, whether computer vision models interpret these images uniformly at a pixel level across different systems remains unclear. Consequently, comparison or replication across systems poses challenges. While several articles report satisfactory performance by deep learning models, validation studies assessing model performance using images from different devices than those used in training are lacking.

Interestingly, despite the Scheimpflug imaging system having a considerable clinical track record in FECD^58^, our review did not uncover any studies attempting to train AI using Scheimpflug image data for FECD management. This is likely due to proprietary closed-source data formats, such as for the Pentacam (OCULUS Optikgeräte GmbH, Germany), which limit bulk export for AI training. Moreover, the newer AS-OCT technology produces higher-resolution cross-sectional images, enabling the differentiation of epithelial and stromal edema—a potentially appealing feature for training deep learning models. Due to the wavelength used in AS-OCT, it generates clearer images for FECD patients with severe edema compared to Scheimpflug tomography, which is prone to be interfered by light scatter. These factors contribute to the preference for training computer vision models using images from AS-OCT over Scheimpflug.

Successful AI model training relies on the availability of ground truth data as a reference standard and a sufficiently large sample size to reveal statistically meaningful patterns. Identifying a detached graft on AS-OCT and delineating endothelial cell edges on specular and confocal microscopy are relatively easy tasks for human eyes, enabling consistent manual labelling by trained experts as ground truth for model training. However, objective detection of corneal edema remains challenging. Corneal thickness is the most widely used surrogate parameter for assessing corneal edema in clinic, though its reliability is limited due to natural variation in normal corneal thickness^59^. Manual labelling of corneal edema on AS-OCT images presents greater challenges than for detecting retinal pathologies like macular edema, where features like intraretinal/subretinal fluid are well-defined as “intraretinal hyporeflective space surrounded by reflective septae” with clear borders, readily recognized by both machines and humans without clinical expertise^60^. In contrast, detecting subclinical corneal edema is challenging, both on slit lamp examination and on quasi-histological OCT images, as early edema in OCT often appears as subtle changes in reflectivity, requiring extensive sub-specialist expertise for accurate delineation. Prognosticating post-corneal transplant graft survival poses equal challenges. Posterior lamellar transplant procedures generally show favorable outcomes. Using AI to uncover the subset of patients at risk of graft failure requires extensive long-term follow-up datasets, which is often unavailable. The lack of a universally accepted reference standard and dataset impede model training in these clinical scenarios.

AI research concerning FECD management has predominantly concentrated on supervised learning approaches like decision trees, regression analysis, and CNNs. Reported performance metrics across these studies exhibit variability, depending on the input data types, AI methodologies, and clinical contexts, making it challenging to compare the performance of different AI algorithms. For classification studies, sensitivity, specificity, and receiver operating characteristic analysis are commonly employed to gauge diagnostic capability. The vast majority of included studies (n = 14), however, did not report other metrics crucial for evaluation and systematic comparison of models’ performance, such as the F1 score or recall. Those are especially key in studies where the training dataset was unbalanced, which is often the case in reports for diseases of low prevalence like FECD. While many classification algorithms exhibited an AUC exceeding 90% when trained and validated on a single dataset, their performance significantly declined when tested on different input datasets^22^, highlighting potential limitations in their clinical application. Dice coefficients have been used to evaluate the alignment between predicted and actual segmentation masks, revealing more variability in segmentation model performance even with data from the same imaging modality. This variability likely stems from the complexity of segmentation tasks, which require precise pixel-level accuracy and detailed boundary delineation, whereas classification tasks involve a broader understanding of overall image features for label assignment.

Validating AI models on datasets distinct from the trained set is crucial to assess the model’s generalizability. While all included studies conducted internal validation, utilizing methods like k-fold cross-validation or random data splitting into training/test sets, only three studies performed external validation^17,22,26^. CNNs, if not appropriately regularized, are susceptible to overfitting, leading to drastic accuracy changes when used with data different from the trained dataset^61^. Interestingly, Bitton et al.^24^, attempting to validate a prior deep learning pipeline using an independent out-of-sample dataset, reported comparable satisfactory performance in line with the original result^23^. This pipeline initially employed a computer vision approach to quantify corneal EF from AS-OCT images and then utilized this value to classify FECD severity. Despite the encouraging classification performance, it became evident that the same EF was not universally applicable, requiring recalibration for different datasets. Studies in other fields have demonstrated that even models exhibiting adequate performance on internal validation may exhibit significant decreases in sensitivity and specificity during external validation^62^, and this decline in performance was evident in the algorithm by Foo et al^26^. Ideally, external datasets should be larger and more representative of the general population^17^.

While computer vision models demonstrate promising performance in various clinical contexts of FECD, their applicability is not always evident across all scenarios. Clinical recognition of advanced FECD with endothelial decompensation is relatively straightforward, graft detachment is visibly discernible on AS-OCT, and assessment of endothelial cells is efficiently performed using manufacture-built software in specular microscopy devices. The development of AI in these areas may offer limited additional utility. Further, it is crucial to consider real-world clinical practices when evaluating AI applications, including the choice of clinical pathway and imaging modalities. For instance, a highly accurate model for diagnosing multiple corneal endothelial diseases may be valuable^17^, but if it relies on a less commonly available and time-consuming imaging modality like IVCM, its practical utility may be constrained. Therefore, research endeavors should address existing clinical care gaps, such as evaluating and quantifying subclinical corneal edema, predicting disease progression, and prognosticating post-corneal transplantation outcomes. Focusing on these aspects is likely to maximize the potential benefits of AI by complementing human limitations, enabling timely interventions to mitigate disease progression and visual loss.

Most of the included studies utilized retrospective data for model development and did not involve masked observers for model performance evaluation, potentially introducing detection bias. The reported studies generally had small sample sizes, with only two studies encompassing more than 500 eyes (including controls), and none of the studies conducted a priori power calculations to estimate the required cohort size. Given that sample size significantly impacts AI algorithm development and the model’s accuracy, further studies with larger sample sizes are warranted.

This systematic review has several limitations. Articles that lacked relevant key terms or were presented solely as abstracts might have been overlooked. However, this rigorous process ensured the inclusion of high-quality articles. Furthermore, the developed pipelines were not always directly comparable. As we intended to be as comprehensive as possible, this review encompassed studies with diverse study designs, sample sizes, input dataset sources, case definitions, imaging modalities, and validation approaches. The heterogeneity among these models posed challenges for meaningful comparisons. Consequently, statistical synthesis and meta-analysis were unfeasible. Additionally, we excluded studies lacking any form of validation and those not conducted on humans, typically proof-of-concept articles outlining experimental steps in model development. This exclusion inevitably resulted in information loss; nevertheless, our intention was to summarise only the approaches with the highest potential for clinical applicability and generalizability.

## Conclusion

We have conducted the most comprehensive review to date on AI algorithms for various clinical contexts of the FECD patient journey. Given the prevalence of FECD as a sight-impairing disease, early detection and prognostication are paramount for effective treatment and to prevent vision loss, making them public health priorities. AI algorithms present promising avenues for achieving these goals and enhancing accessibility. Our summary of relevant publications focuses on input data, algorithm selection, and validation approaches. Future research could explore the potential of AI algorithms based on derived topographic, keratometric and aberrometric parameters rather than only the raw images, which may be less computationally intensive. Additionally, considering known risk factors like older age, female sex, a white ethnicity, and the genetic heterogeneity of FECD^1^, a multimodal approach that merges information from corneal images with demographic and genetic data should be explored. This review offers critical insights for researchers, clinicians, and policymakers, aiding their understanding of existing research and guiding future health service strategies.

## Data Availability

All data produced in the present work are contained in the manuscript

---

**Supplementary Table 1:**
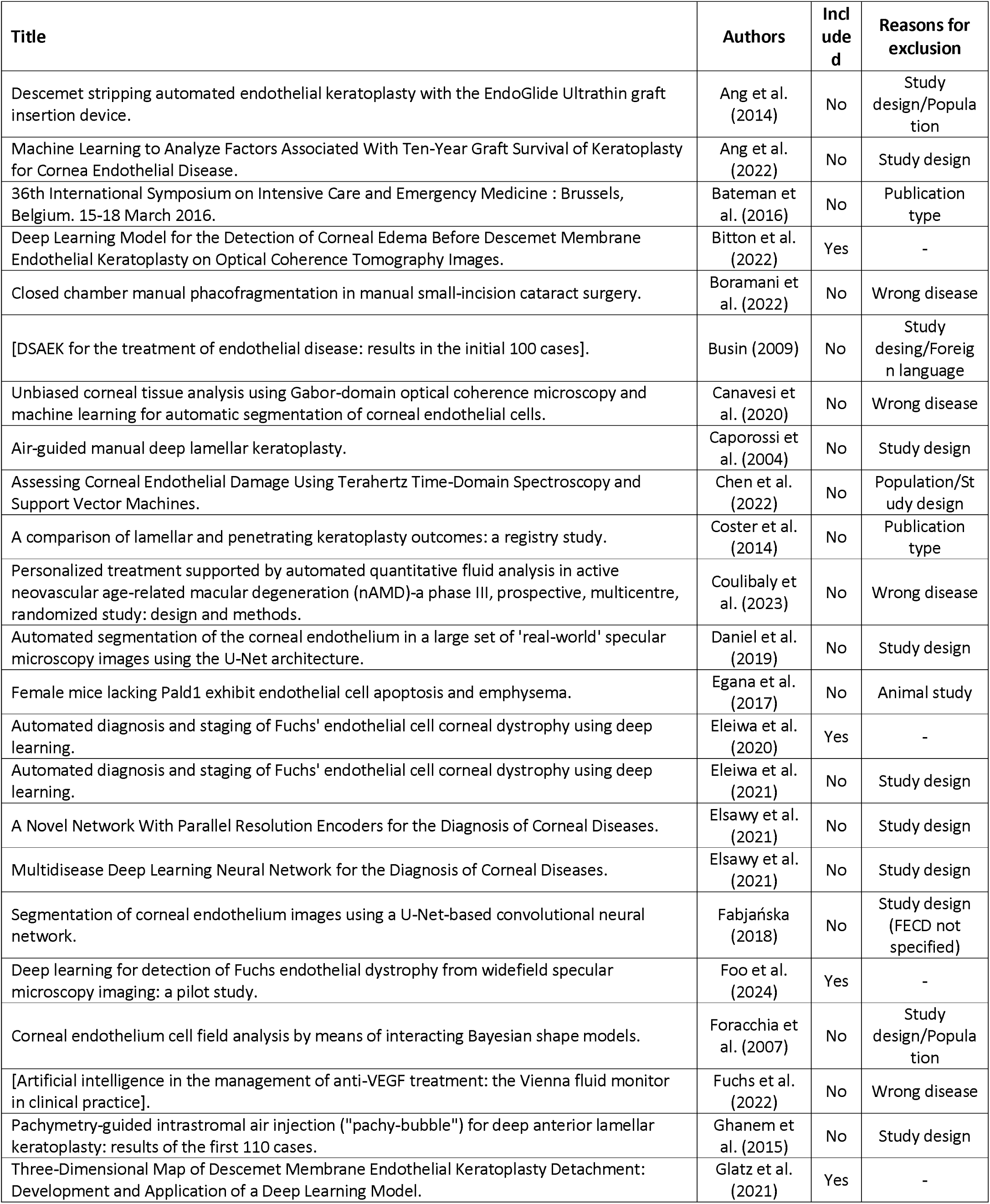
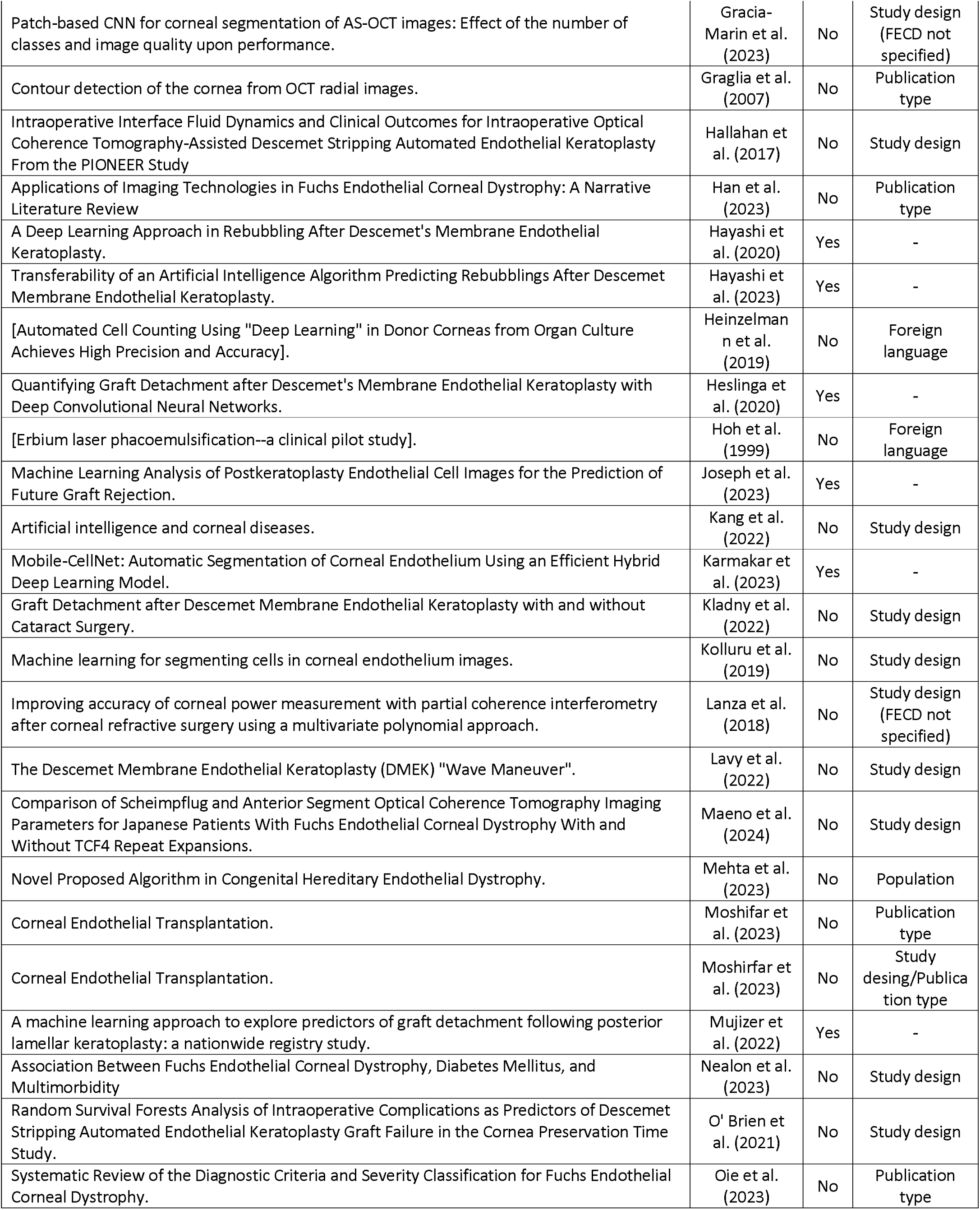
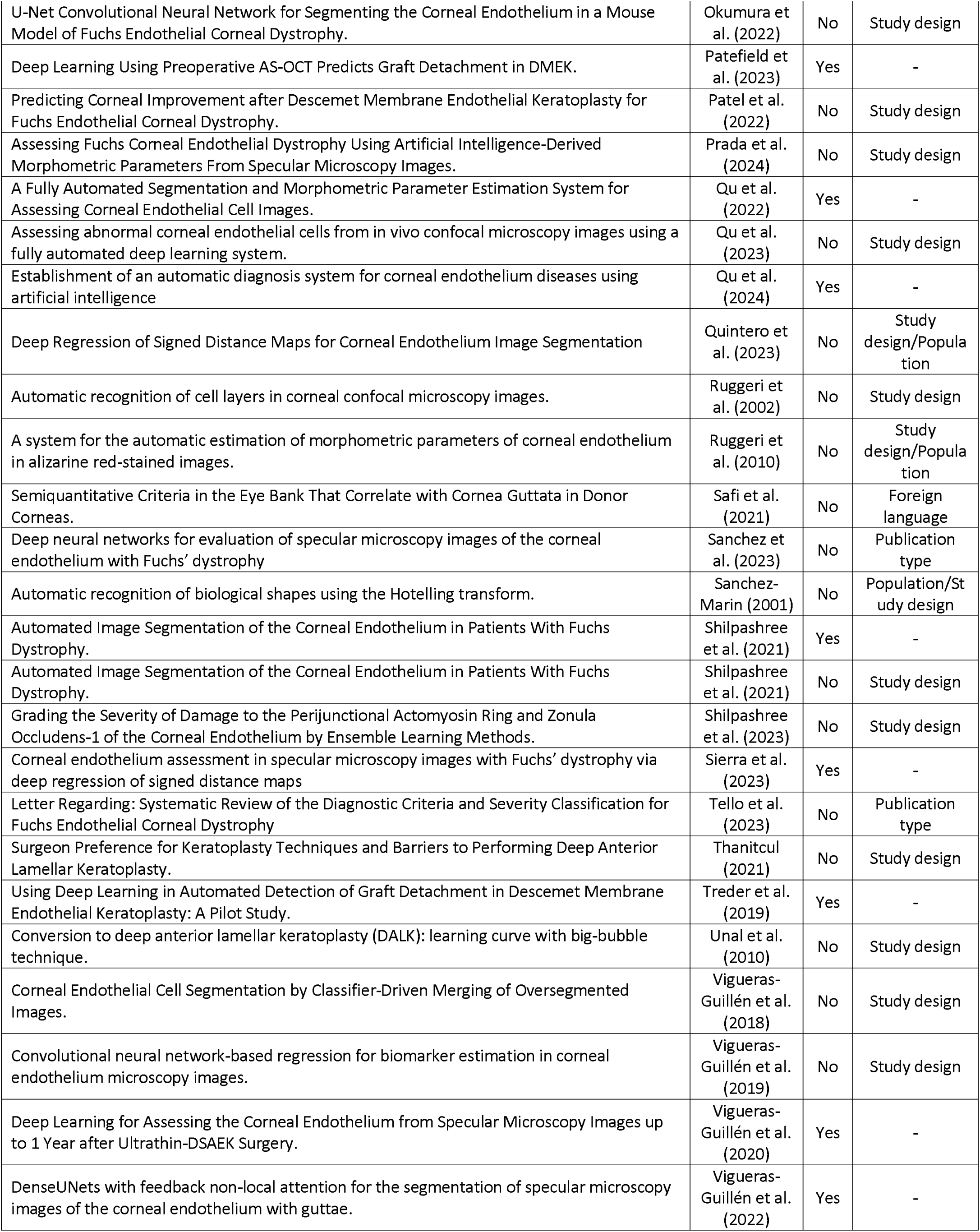
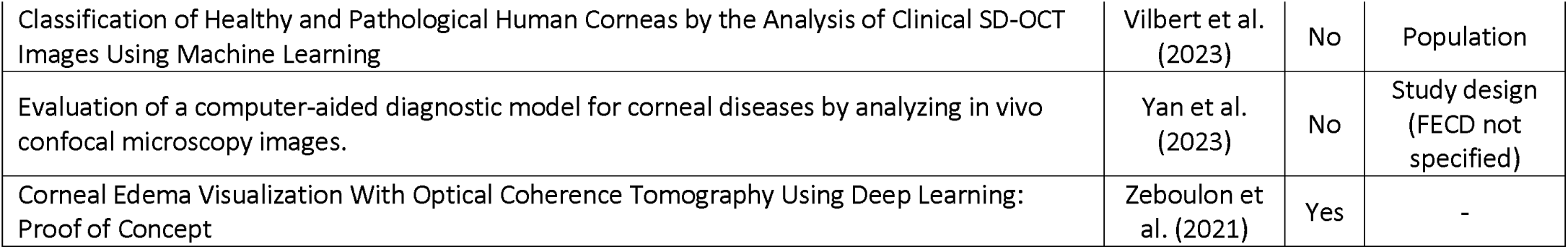
Summary of the published studies reviewed for inclusion, indicating whether each study was included or excluded, along with the reason for exclusion when applicable.

**Supplementary Table 2:**
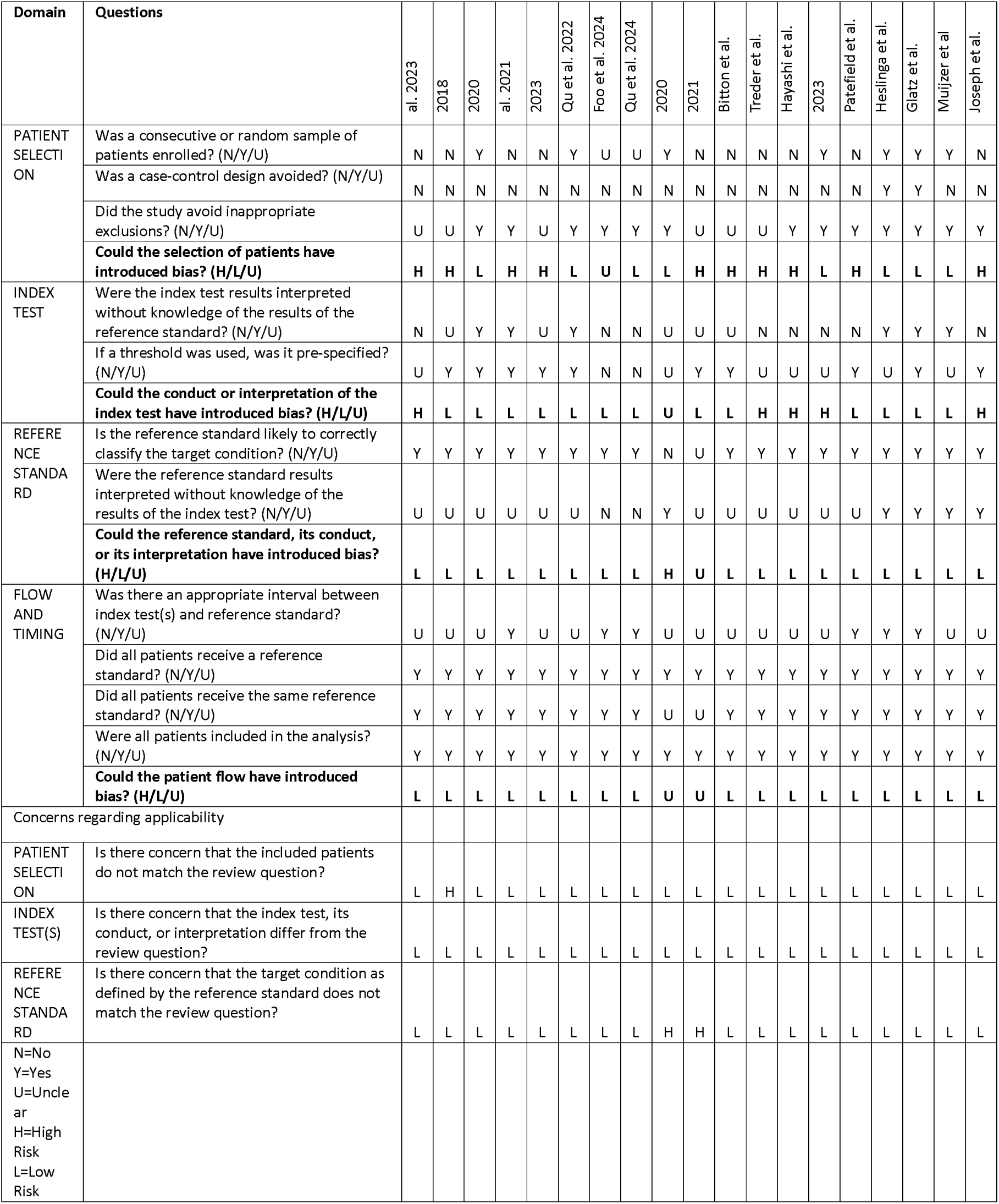
Results of the QUADAS (Quality Assessment of Diagnostic Accuracy Studies)-2 bias assessment tool for included studies.

